# Mutations in TP53 and other heterogeneous genes help to distinguish metastases from new primary malignancies

**DOI:** 10.1101/2020.11.11.20226845

**Authors:** Owen W. J. Prall, Ravikiran Vedururu, Peter Lau, Andrew Fellowes

**Affiliations:** Departments of Pathology, Peter MacCallum Cancer Centre, East Melbourne, VIC 3002, Australia; Cancer Medicine, Peter MacCallum Cancer Centre, East Melbourne, VIC 3002, Australia

**Keywords:** mutation profile, mutation diversity, TP53, metastasis

## Abstract

**Background:** Distinguishing metastases from new primary malignancies or vice versa is important because misclassification can result in inappropriate management. However, for some cases this distinction can be challenging, particularly for squamous cell carcinomas in which the usual surgical pathology approach, predominantly morphology and immunohistochemistry, are frequently non-contributory. We analysed tumor-associated mutations in order to determine whether they could help with this diagnostic dilemma.

**Methods:** Mutations in specific genes were identified with cBioPortal, a large publically available tumor sequence data set. Genes were selected based upon either their high overall prevalence of mutation, or their inclusion in an in-house tumor sequencing set. Tumor types analysed included various common adenocarcinomas, squamous cell carcinomas from multiple sites, urothelial carcinoma and melanoma. Individual mutations and sets of mutations within gene cohorts were compared by their diversity (or heterogeneity) index, prevalence and cumulative prevalence. We demonstrated the utility of this method by performing in-house sequencing of candidate genes in tumors from three patients for which morphology and immunohistochemistry were unable to distinguish between a metastasis and a new primary malignancy.

**Results:** Sequence data from relatively small cohorts of candidate genes readily identified highly diverse, low prevalence mutation profiles in most common malignancies including squamous cell carcinomas. The diversity index predicted the likelihood of an identical mutation profile occurring in an unrelated tumor. High yield gene cohorts could be predicted based on the primary tumor type. Most cohorts included TP53 due to both its high mutation prevalence and high mutation index of diversity. Identical, low prevalence mutations in multiple tumors from patients in the three case studies provided strong diagnostic certainty for metastases rather than new primary malignancies.

**Conclusions:** Most common tumors, including squamous cell carcinomas, have a readily identifiable mutation profiles that occur at a sufficiently low prevalence to effectively barcode or fingerprint the tumor. An identical mutation profile in a primary tumor and a new lesion provides strong evidence for a metastasis and effectively excludes a new primary malignancy, providing diagnostic confidence and aiding clinical management certainty.

## BACKGROUND

Advances in screening and surveillance of cancer patients have resulted in the increased detection of metastases. Along with increased patient age, these have also resulted in increased detection of new primary malignancies. Although radiological and pathological data may help to distinguish new primary malignancies from metastases of previous tumors, for a substantial proportion of cases this is not possible, resulting in considerable clinical uncertainty. Misdiagnosis of a new primary malignancy as a metastasis, or vice versa, results in incorrect staging and likely inappropriate management.

The behaviour, morphology and marker expression of tumors results from their genetic and epigenetic status. Genetic mutations in tumors are mainly sought by oncologists for determining suitability to targeted therapies. They are relatively infrequently used by pathologists to aid diagnosis. Many critical or driver mutations occur early within the history of the tumor, and although other mutations typically accumulate during tumor progression, most driver mutations are typically maintained, particularly in the absence of a targeted therapy. Detection of some types of tumor mutations (e.g. missense, nonsense, small insertions/deletions) has recently become relatively cost-effective and rapid due to advance in DNA sequencing. This has enabled comprehensive sets of cancer exomes to be generated, many of which are now freely and publically available, as well as reliable detection of these mutations in individual patient tumor samples.

Data shown here indicates that most primary malignancies have mutation profiles that are of sufficiently low frequency to “barcode” and track subsequent metastases. Surprisingly, relatively few genes are necessary to provide useful quantitative data to help distinguish a metastasis from a second primary tumor. Three cases studies are provided to illustrate this point. Readily available database sequence information was used to predict other genes that will be useful for this purpose in other typically problematic tumors including squamous cell carcinoma.

## METHODS

### Public database analysis

cBioPortal [1, 2] was queried for non-synonymous mutations in specific genes. Large published exome sequence datasets from common tumor types were analysed including 1261 breast carcinomas [3-6], 55 cholangiocarcinomas [7, 8], 365 colorectal adenocarcinomas [9-11], 248 uterine endometrioid carcinomas [12], 146 esophageal adenocarcinomas [13], 32 gallbladder adenocarcinomas [14], 576 lung adenocarcinomas [15-18], 316 ovarian serous adenocarcinomas [19], 208 pancreatic adenocarcinomas [20, 21], 759 prostatic adenocarcinomas [22-26], 447 gastric adenocarcinomas [27-30], 279 squamous cell carcinomas (SCC) from head and neck [31, 32], 178 lung SCCs [33], 225 esophageal SCCs [34, 35], 29 metastatic skin SCCs [36], 376 bladder urothelial carcinomas [37-40] and 212 melanomas [41, 42].

Microsoft Excel pivot tables were used to determine the prevalence of mutations in individual genes, and in combinations of genes. The expected prevalence of two different mutations was calculated as the product of their individual prevalence. The diversity, or heterogeneity, of sequences was quantified by Simpson’s index D, where 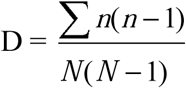 [43] and was expressed as the index of diversity, 1-D [44]. In this equation n = the number of tumors with each sequence combination, and N = number of all tumors in the set. The index of diversity was termed the “overall heterogeneity” for the set of all tumors (i.e. including those with wild type sequences). When only tumors with mutated sequences were considered the index of diversity was termed the “mutation-specific heterogeneity”.

### In house next generation sequencing

For the three case studies formalin-fixed paraffin-embedded (FFPE) material was enriched for tumor cells via microdissection. DNA was extracted using either the DNeasy Blood and Tissue Kit or EZ1 DNA Tissue Kit (Qiagen) according tomanufacturer’s instructions. DNA quality was assessed by an in-house designed qPCR assay based on the Ct values of 100bp and 300bp amplicons of non-cancer related genes.

Library enrichment was performed by multiplex PCR on 48×48 Access Array Microfluidics chips (Fluidigm) followed by sequencing of a 138 amplicon panel. This panel was designed in-house to be FFPE-compatible and targets mutation hotspots in 19 genes with prognostic and/or therapeutic relevance (Table 1). Where possible, mutation hotspots were covered with multiple redundant amplicons. Data was analysed via an in-house developed pipeline. Nucleotide sequences in FASTQ format were demultiplexed with bcl2fastq Conversion Software v1.8.4 (Illumina). Sequence alignment was performed with the Needleman–Wunsch algorithm, and variant calling with Varscan2.

**Table 1.** In house sequencing. The in house 138 amplicon panel targets exons from 19 genes with prognostic and/or therapeutic relevance.

| **Gene** | **Refseq ID** | **Exons covered** | **Hotspot codons** |
| --- | --- | --- | --- |
| **AKT1** | NM_005163.2 | 3 | 17 |
| **ALK** | NM_004304.4 | 20, 22, 23, 24, 25 | 1174, 1245, 1275 |
| **BRAF** | NM_004333.4 | 11, 15 | 444, 469, 600 |
| **CDKN2A** | NM_000077.4 | 1, 2 | 58, 80 |
| **EGFR** | NM_005228.3 | 18, 19, 20, 21 | Multiple |
| **FGFR2** | NM_000141.4 | 7, 9, 12 | 252, 382, 549 |
| **FGFR3** | NM_000142.4 | 7, 10, 16 | 249, 380, 716 |
| **HER2** | NM_001005862.2 | 8, 17, 19, 20,<br>21, 22 , 27 | 310, 655, 678, 755, 842 |
| **KIT** | NM_000222.2 | 9, 11, 13, 17 | Multiple |
| **KRAS** | NM_033360.2 | 2, 3, 4 | Multiple |
| **MAP2K1** | NM_002755.3 | 2, 3 | 53-57, 124-130 |
| **MET** | NM_000245.2 | 14 | 3' splice site mutations |
| **NRAS** | NM_002524.3 | 2, 3, 4 | Multiple |
| **PDGFRA** | NM_006206.4 | 12, 14, 18 | Multiple |
| **PIK3CA** | NM_006218.2 | 10, 21 | 542, 545, 1047 |
| **PTEN** | NM_000314.4 | All | Multiple |
| **RAC1** | NM_006908.4 | 2 | 29 |
| **RNF43** | NM_017763.4 | 3, 9 | 117, 659 |
| **TP53** | NM_000546.5 | All | Multiple |

## RESULTS

### Low prevalence mutation profiles are present in many tumors

The prevalence of mutations was assessed in an eight-gene subset (termed cohort-8) from the 19 genes that are frequently sequenced for prognostic and predictive data at our institution. Cohort-8 included TP53, KRAS and BRAF as above, as well as AKT1, NRAS, PDGFRA, KIT and PIK3CA. Data from 3753 malignancies was analysed with cBioPortal [1, 2]. These were predominantly adenocarcinomas from breast, colorectum, endometrium (endometrioid type), lung, ovary (serous type), pancreas, thyroid (papillary type) and prostate, as well as melanomas. Overall, the proportion of tumors mutated for each gene was similar to well established data (Fig.1A).

**Figure 1.**
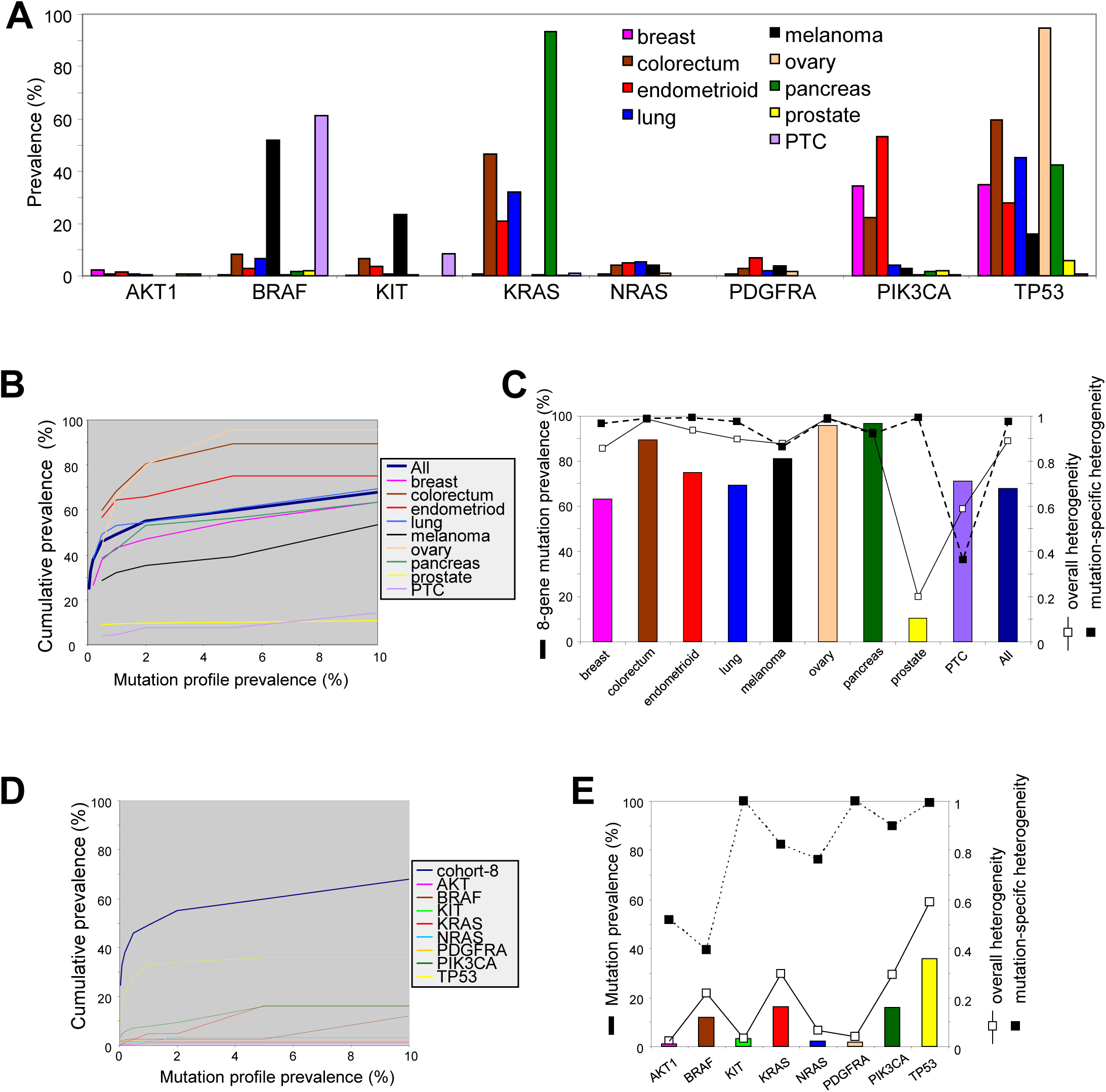
Heterogeneity of cohort-8 in adenocarcinomas and melanoma. **A**. Prevalence of mutations in each gene in the cohort-8. **B**. Cumulative prevalence of mutation profiles for cohort-8. Where possible, the cumulative prevalence was calculated for individual mutation profile prevalences from 0.05 to 10%. **C**. Mutation prevalence, overall heterogeneity and mutation-specific heterogeneity of cohort-8. **D**. Prevalence of mutation profiles for each gene in cohort-8 in adenocarcinomas and melanoma. Where possible, the cumulative prevalence was calculated for individual mutation profile prevalences from 0.05 to 10%. **E**. Cumulative prevalence, overall heterogeneity and mutation-specific heterogeneity for each gene in cohort-8.

Mutation profiles in cohort-8 were generally useful to distinguish metastases from new primary tumors. For example, 55.1% of all tumors had a mutation profiles in cohort-8 with a prevalence of <2%, (Fig.1B), in other words occurring in less than 1 in 50 other tumors. Furthermore, 45.9% of tumors had cohort-8 mutation profile prevalences of <0.5%, i.e. occurring in <1/200 other tumors (Fig.1B). There was substantial variation between different tumor types in the proportion of tumors that had low prevalence mutation profiles. The proportion was very high for ovarian and colorectal carcinomas in which about 80% of tumors had a mutation profile prevalence <2%, but was very low for prostatic and papillary thyroid carcinomas in which <15% of tumors had a mutation profile prevalence <10% (Fig.1B).

The proportion of tumors with low prevalence mutation profiles was a consequence of the overall heterogeneity of genes in cohort-8. Simpson’s index of diversity [43, 44] was used to quantify heterogeneity. This index expresses the probability that two randomly selected tumors will have different mutation profiles. An index of diversity of 1 represents infinite heterogeneity, and an index of 0 represents no heterogeneity. Overall heterogeneity is contributed to by both the prevalence of mutations and mutation-specific heterogeneity. A set of tumors dominated by only one or two mutation profiles will be less heterogeneous than a set composed of numerous different mutation profiles. The heterogeneity of cohort-8 was high in tumor subtypes that had abundant low prevalence mutations (compare Fig.1B and 1C). For example, heterogeneity was >98% for ovarian and colorectal carcinomas in which cohort-8 showed both highly prevalent and highly heterogeneous mutations. Tumors with only highly prevalent mutations (e.g. papillary thyroid carcinoma) or highly heterogeneous mutations (e.g. prostate) in cohort-8, but not vice versa, had a low overall tumor heterogeneity (20% for prostate carcinoma and 60% for papillary thyroid carcinoma).

### TP53 heterogeneity

When all 3753 malignancies were considered together, the greatest contribution to the heterogeneity of cohort-8 was from TP53. TP53 was the most frequently mutated gene (prevalence of 35.8%) and had the most diverse set of mutations (mutation-specific heterogeneity of 0.99) resulting in the highest gene diversity (overall heterogeneity of 0.58). Over 98% of individual TP53 mutations had prevalences of <2%, and about a third of tumors (32.9%) had a TP53 mutation with a prevalence of <1%. Mutations were widely distributed throughout the TP53 sequence with some low amplitude hotspots (Fig.2C), as previously described [45]. The mean prevalence of TP53 mutations ranged from 0.25% (prostate, standard deviation of 0.06%) to 0.63% (pancreas, standard deviation of 0.44%). The commonest TP53 mutation overall was R175H, with an overall prevalence of 1.73% and the only mutation to have a prevalence of >3.5% in a specific tumor subtype (7.4% of colorectal adenocarcinomas, Fig.2C). The mutation-specific heterogeneity of KIT and PDGFRA was even greater than for TP53, 1.0 and 0.9997, respectively (Fig.1E), as all, or almost all mutations in these genes were unique. However, mutations were rare for both KIT and PDGFRA (prevalence of 3.7% and 1.6%, respectively) resulting in low overall heterogeneity (0.032 and 0.041), as this analysis includes wild-type sequences. The prevalence of BRAF mutations was relatively high at 12% (Fig.1E). However, 78% of all BRAF mutations were V600E, which resulted in a low overall heterogeneity (overall index of diversity of 0.22). AKT had the lowest overall heterogeneity (0.023) due to both low mutation prevalence (1.15%) and low mutation-specific heterogeneity (0.52). 70% of all AKT mutations were E17K.

**Figure 2.**
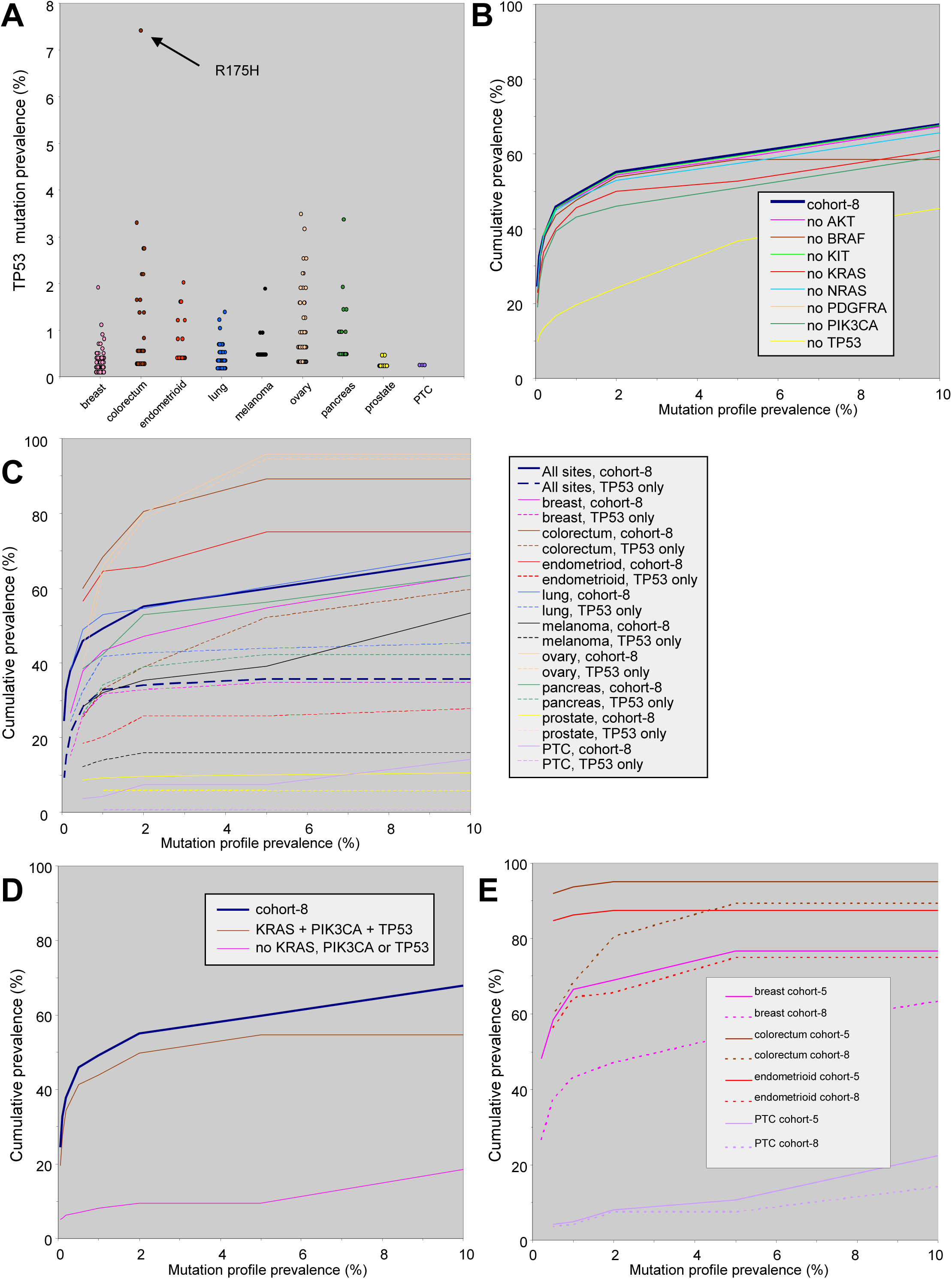
The contribution of specific genes to mutation profile heterogeneity. **A**. Scatter plot of the prevalence of individual mutations in TP53 in specific tumor types. **B**. Cumulative prevalence of mutation profiles for cohort-8 in all tumors combined after each individual gene was removed. **C**. Cumulative prevalence of mutation profiles for TP53 alone versus cohort-8. **D**. Cumulative prevalence of mutation profiles for the KRAS, PIK3CA, TP53 cohort versus the remaining 5 genes from cohort-8 (i.e. without KRAS, PIK3CA and TP53). **E**. Cumulative prevalence of mutation profiles for cohort-5 (as predicted from Table 2) versus cohort-8. Tumors are as indicated. Where possible, the cumulative prevalence was calculated for individual mutation profile prevalences from 0.05 to 10%.

In about a third of all tumors (34.1%) sequencing TP53 alone identified a mutation profile that would occur in <1/50 other tumors (prevalence of <2%, Fig.1D). There were also important but relatively minor contributions to the overall heterogeneity of cohort-8 from PIK3CA and KRAS, for which a mutation profile prevalence of <2% was present in 9.4% and 4.8% of all tumors, respectively (Fig.1D). A mutation profile prevalence of <2% was only present in 24.1% of tumors when TP53 was removed, compared to 55.1% when TP53 was included (Fig.2B). TP53 by itself provided a useful low prevalence mutation profile in most tumor types (Fig.2C). For colorectal adenocarcinomas the mutation profile prevalence of TP53 alone was similar to that for all other seven genes together (not shown). However, TP53 was less useful for endometrioid and papillary thyroid carcinomas, and melanoma (Fig.2C). This was due to low prevalence rather than low mutation-specific heterogeneity (not shown). When all the malignancies were considered together, the three genes with the greatest heterogeneity (TP53, PIK3CA, KRAS, Fig.1E) together provided a mutation profile prevalence nearly equivalent to cohort-8 (Fig.2D).

### Identifying other heterogeneous genes in adenocarcinomas

In order to improve mutation profiling, cBioPortal data from common tumor types was used to identify genes with frequent sequencing-detectable mutations (Table 2). For many tumor types most of these genes have been shown to be drivers of tumorigenesis and therefore likely maintained in metastases. TP53 remained one of the most heterogeneous genes in many tumor types, with exception of endometrioid and papillary thyroid carcinoma and melanoma

**Table 2.**
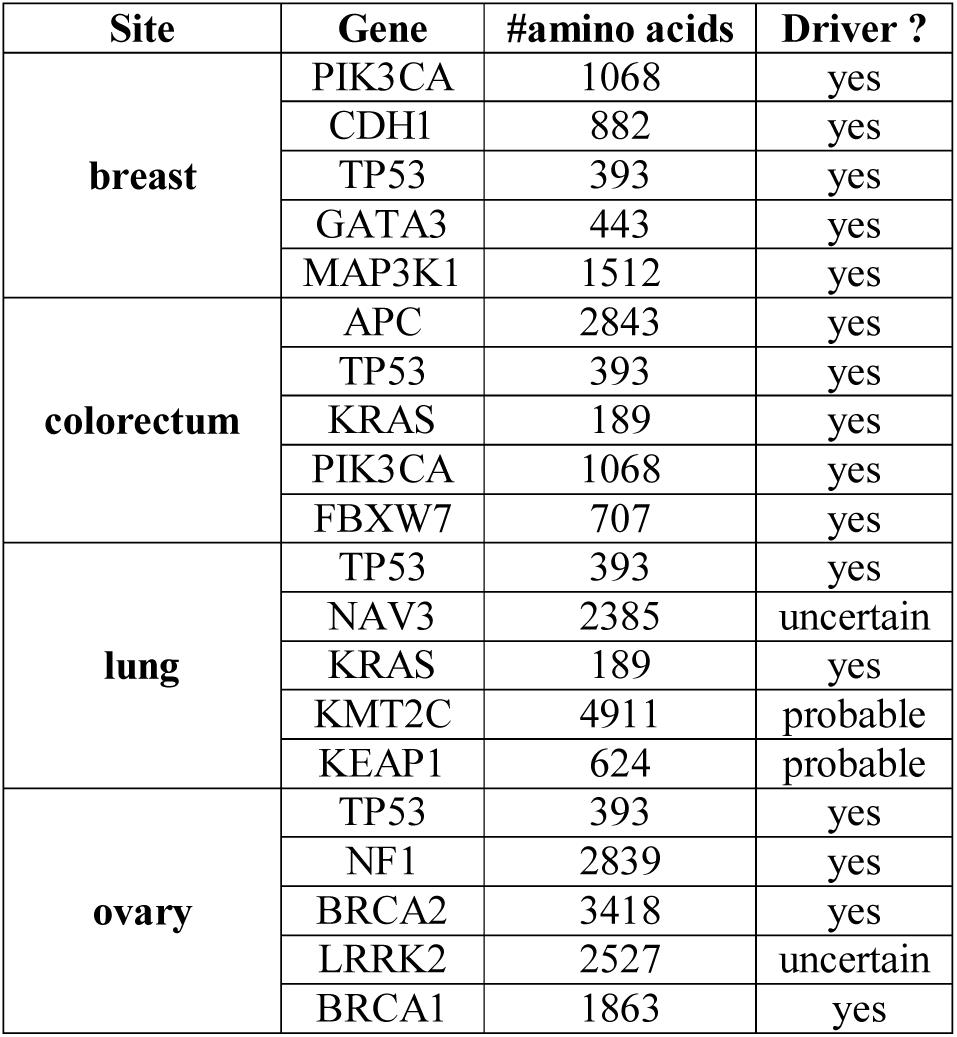
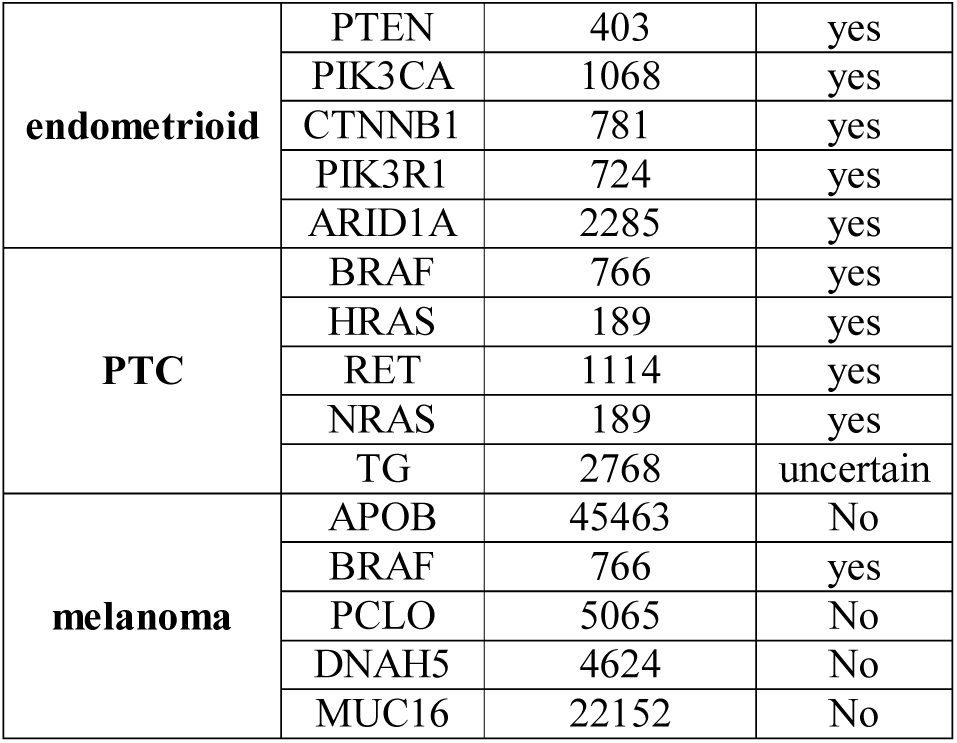
Frequently mutated genes in adenocarcinoma and melanoma. Five of the most frequently mutated genes in adenocarcinomas from various sites and melanoma predicted from cBioPortal data. Protein length (amino acids, aa) and tumor driver status are indicated.

Where possible, a new cohort for each tumor subtype was constructed from this data (cohort-5) and compared to cohort-8. Heterogeneous driver genes with large coding sequences were frequently identified in ovarian and lung adenocarcinoma. Large size likely reduced the success of reliably obtaining their sequence. Also, for melanoma four of the top five most frequently mutated genes had no known role in neoplasia. They are likely UV-induced passenger mutations that accumulate in part due of their large size. Genes of uncertain driver status were also relatively common in lung adenocarcinoma. Therefore ovarian and lung adenocarcinomas and melanomas and were excluded from further analysis, although for serous ovarian carcinoma the predictive value of the original 8-gene cohort was already very high and unlikely to be improved.

For most adenocarcinomas low prevalence mutation profiles were more easily identified with cohort-5 compared to cohort-8 (Fig.2E). For example, a mutation profile prevalence of <2% was present in 95.1% of colorectal carcinomas with cohort-5, compared to in 80.0% of tumors with cohort-8. The exception was papillary thyroid carcinoma in which a mutation profile prevalence of <2% was present in only about 8% of tumors using either cohort-5 or cohort-8 Sequence-detected mutations were highly prevalent in papillary thyroid carcinoma (80.5%), but their diversity remained low as most remained restricted to BRAF V600E (58.1%) and the new genes in cohort-5 added very little to the overall heterogeneity.

### Identifying heterogeneous genes in squamous cell carcinomas

Distinguishing between poorly differentiated carcinomas with either squamous morphology or p63/p40 expression is generally more difficult than for adenocarcinomas. The differential diagnosis includes squamous cell carcinoma (SCC), urothelial carcinoma, and sometimes other carcinomas (e.g. salivary gland, metaplastic breast). It can be very difficult to separate primary versus metastatic SCC. Frequently mutated genes common to SCCs from head and neck, lung, skin and esophagus, and urothelial carcinoma were identified from cBioPortal data in order to develop a useful mutation profiling cohort. As above, genes of unproven driver status or with large coding sequences were excluded from further analysis. This resulted in a 3-gene subset (cohort-SCC) composed of TP53, CDKN2A and PIK3CA. One of these three genes was mutated in >72% of SCCs (Fig.3A). The overall diversity of cohort-SCC was high, with a heterogeneity index of >0.92 in all SCCs due to a very high mutation-specific heterogeneity (>0.99, Fig.3A). Mutation-specific heterogeneity of cohort-SCC was also very high in urothelial carcinomas (0.99), although the prevalence of mutation was lower than for SCCs (53%), resulting in a lower overall heterogeneity (index of 0.78). Again, TP53 was the most frequently mutated of the three genes in both SCC and urothelial carcinoma (Fig.3B). The mean prevalence for individual TP53 mutations ranged between 0.49% (skin SCC, standard deviation = 0.13%) to 0.67% (urothelial carcinoma, standard deviation = 0.58%, Fig.3C). The commonest TP53 mutation overall was R248W, with a prevalence of 4.0% in urothelial carcinoma (Fig.3C). Clinically useful data is generated with cohort-SCC. A mutation profile prevalence of <2% was present in at least 72% of SCCs from most sites (there were too few skin SCCs to calculate this figure) and a prevalence of <5% (i.e. occurring by chance in 1/20 different SCCs) was present in 72% of esophageal SCCs, >82% of SCCs from the other sites, and 53% of urothelial carcinomas (Fig.3D).

**Figure 3.**
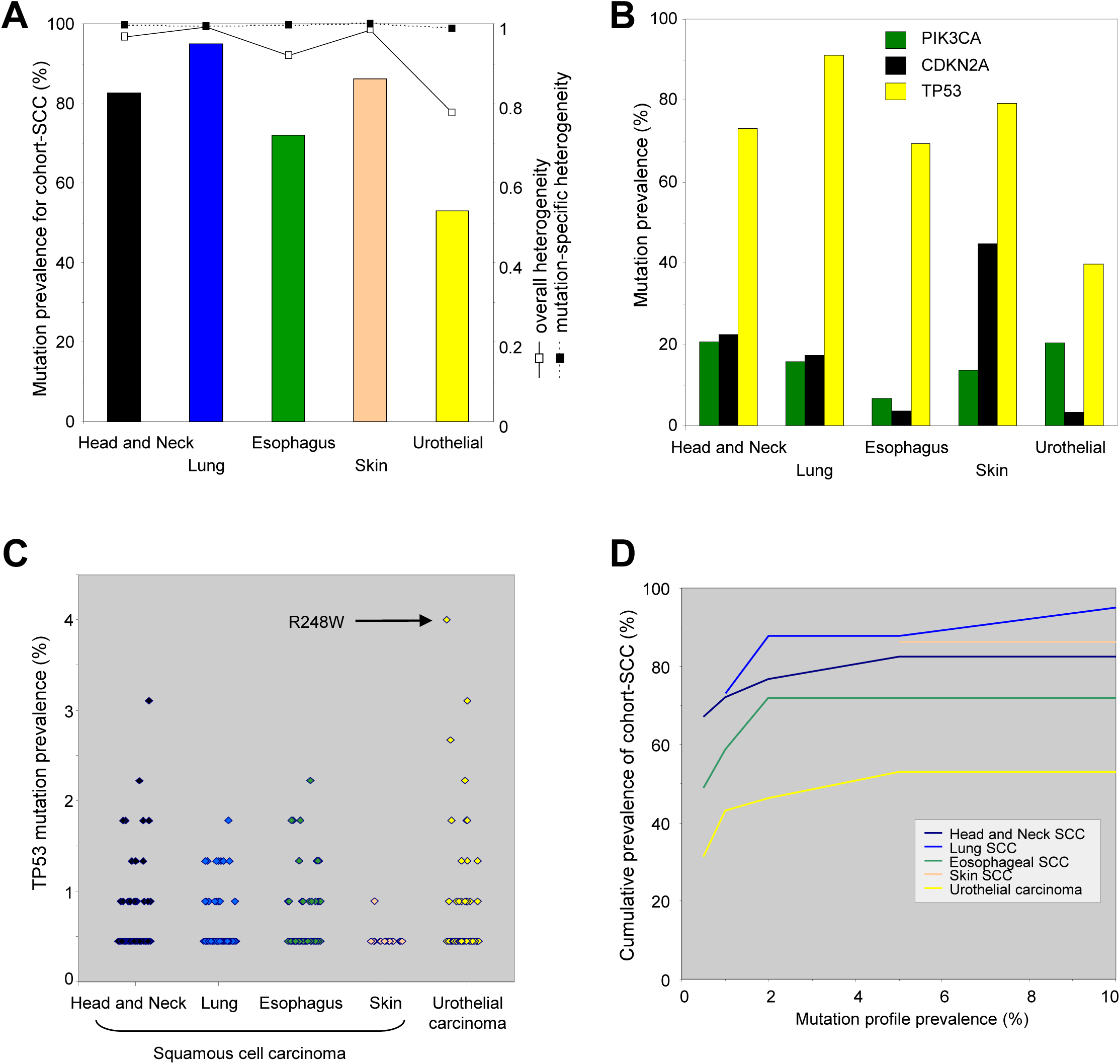
Heterogeneous genes in squamous cell carcinomas. **A**. Mutation prevalence, overall heterogeneity and mutation-specific heterogeneity for cohort-SCC (TP53, PIK3CA and CDKN2A) in squamous cell carcinomas and urothelial carcinoma. **B**. Prevalence of mutations of each gene in cohort-SCC in squamous cell carcinomas and urothelial carcinoma. **C**. Scatter plot showing the prevalence of individual mutations in TP53 for each tumor type. **D**. Cumulative prevalence of mutation profiles in cohort-SCC in squamous cell carcinomas and urothelial carcinoma.

### Case histories

The bioinformatics analysis above suggests mutations of sufficiently low prevalence can effectively “barcode” primary malignancies and track their metastases. The usefulness of this method is illustrated by 3 case studies.

### Case 1

A male in his 50’s presented with stage 3A rectal adenocarcinoma and was treated with neoadjuvant chemo-radiotherapy and anterior resection. He remained disease-free for four years until a 7mm left lower lobe nodule and multiple suspicious lymph nodes (left pulmonary hilar, subcarinal, left lower paratracheal) were detected by surveillance imaging. FNA of the subcarinal and paratracheal lymph nodes showed adenocarcinoma. The left lower lobe nodule and nodes were surgically excised. Both were invaded by a tumor expressing TTF1 (both clones SP141 and 8G7G3/1, Fig.4B and not shown). However, tumor in the resection specimens and FNA also had an enteric morphology (Fig.4A) and immunophenotype (CK7-CK20+ CDX2+, not shown). The original rectal tumor was reviewed. It was morphologically similar to the lung and nodal tumors, but was TTF1-negative. The lung and nodal tumors were thought to be most likely metastases from the rectum but their distribution and TTF1 expression raised the possibility of a primary lung adenocarcinoma. In house sequencing showed the same point mutations in TP53 (844C>T, R282W) and KRAS (183A>T, Q61H) in the rectal, lung and subcarinal lymph node tumors. The normal lung parenchyma was wild-type for TP53 (and KRAS), arguing against germline mutation.

**Figure 4.**
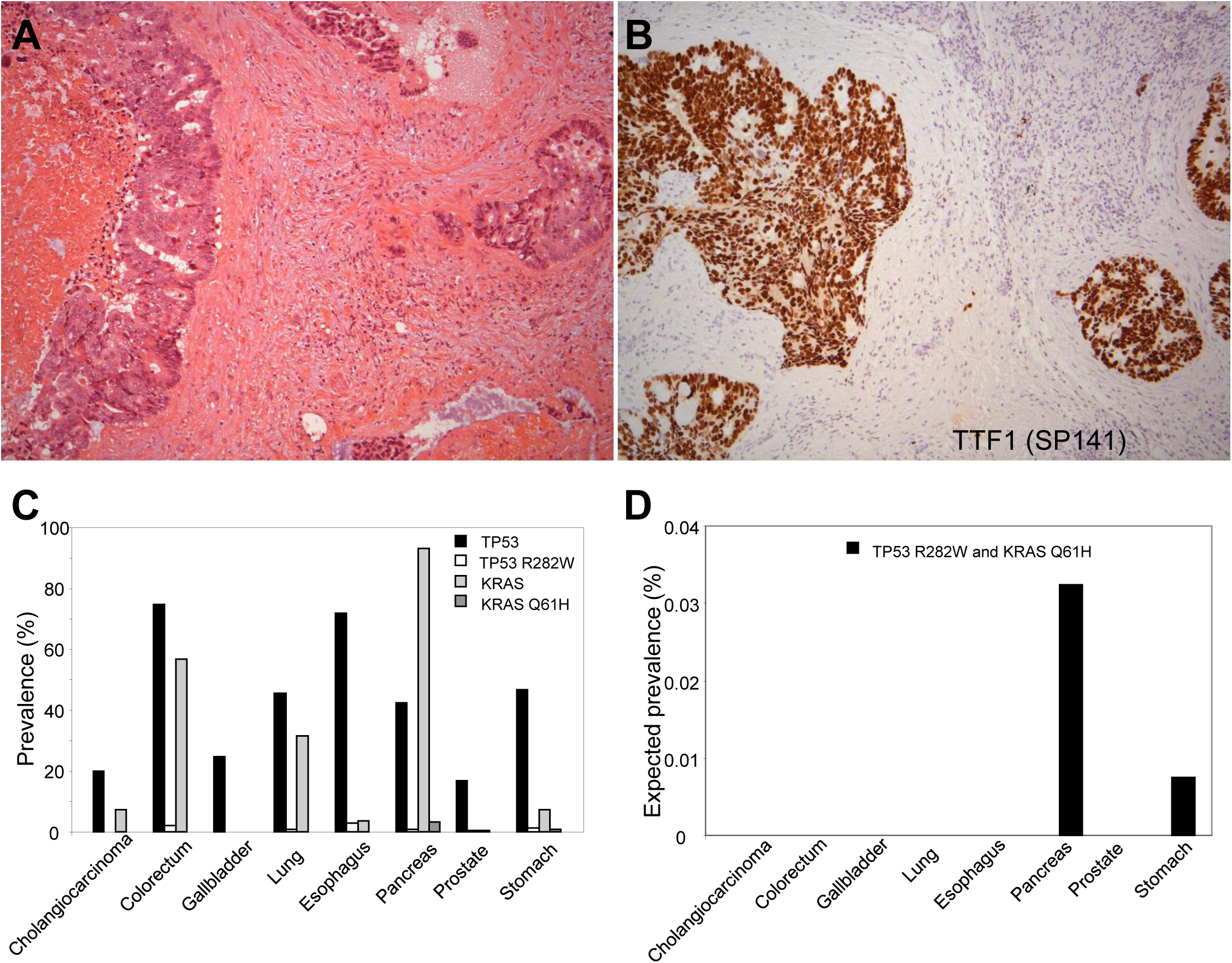
Case study 1. **A, B**. Left lower paratracheal lymph node tumor morphology by H&E section (A) and expression of TTF1 by immunohistochemistry (B). **C**. Prevalence of all sequence-detected TP53 and KRAS mutations, and specific TP53 (R282W) and KRAS (Q61H) mutations in adenocarcinomas from multiple sites. Data from cBioPortal (see text). **D**. Expected prevalence of the co-occurrence of TP53 R282W and KRAS Q61H mutations in adenocarcinomas from various sites.

Analysis of cBioPortal mutation data [1, 2] from 2549 adenocarcinomas relevant to males in showed that while mutation of either TP53 or KRAS was common from colorectal, lung and other sites, TP53 R282W or KRAS Q61H were relatively rare (Fig.4C). The combination of TP53 R282W with KRAS Q61H was not identified in any of adenocarcinomas including 365 colorectal adenocarcinomas. This was not surprising considered that the combination of TP53 R282W and KRAS Q61H, if independent, was predicted to present in <0.035% for any adenocarcinoma subtype (Fig.4D). The low prevalence of the TP53 R282W and KRAS Q61H combination strongly supported the diagnosis of metastatic rectal adenocarcinoma over a new primary lung adenocarcinoma.

### Case 2

A female in her 30’s presented with numerous liver metastases. An avid transverse colon lesion was detected by FDG-PET but was not evident on CT imaging. A diagnostic laparoscopy was converted to an open right hemicolectomy and omentectomy. Histology showed a poorly differentiated tumor invading the colonic wall (Fig.5A) which in immunostains was pan-cytokeratin+ (Fig.5B) and CK7-CK20+ CDX2+ (not shown) consistent with a primary colonic adenocarcinoma. There was also extensive undifferentiated omental tumor that raised the possibility of a second malignancy (Fig.5C). The omental tumor was negative for numerous cytokeratins (Fig.5D), as well for CDX2 and numerous markers of melanocytic and haematolymphoid differentiation (not shown). Mismatch repair protein expression was preserved in both components. In house DNA equencing showed that the colonic adenocarcinoma and undifferentiated omental tumor both had the same point mutations in TP53 (524G>A, R175H) and BRAF (1799T>A, V600E). An incidental serrated colonic polyp from the patient was wildtype for TP53 arguing against a germline mutation.

**Figure 5.**
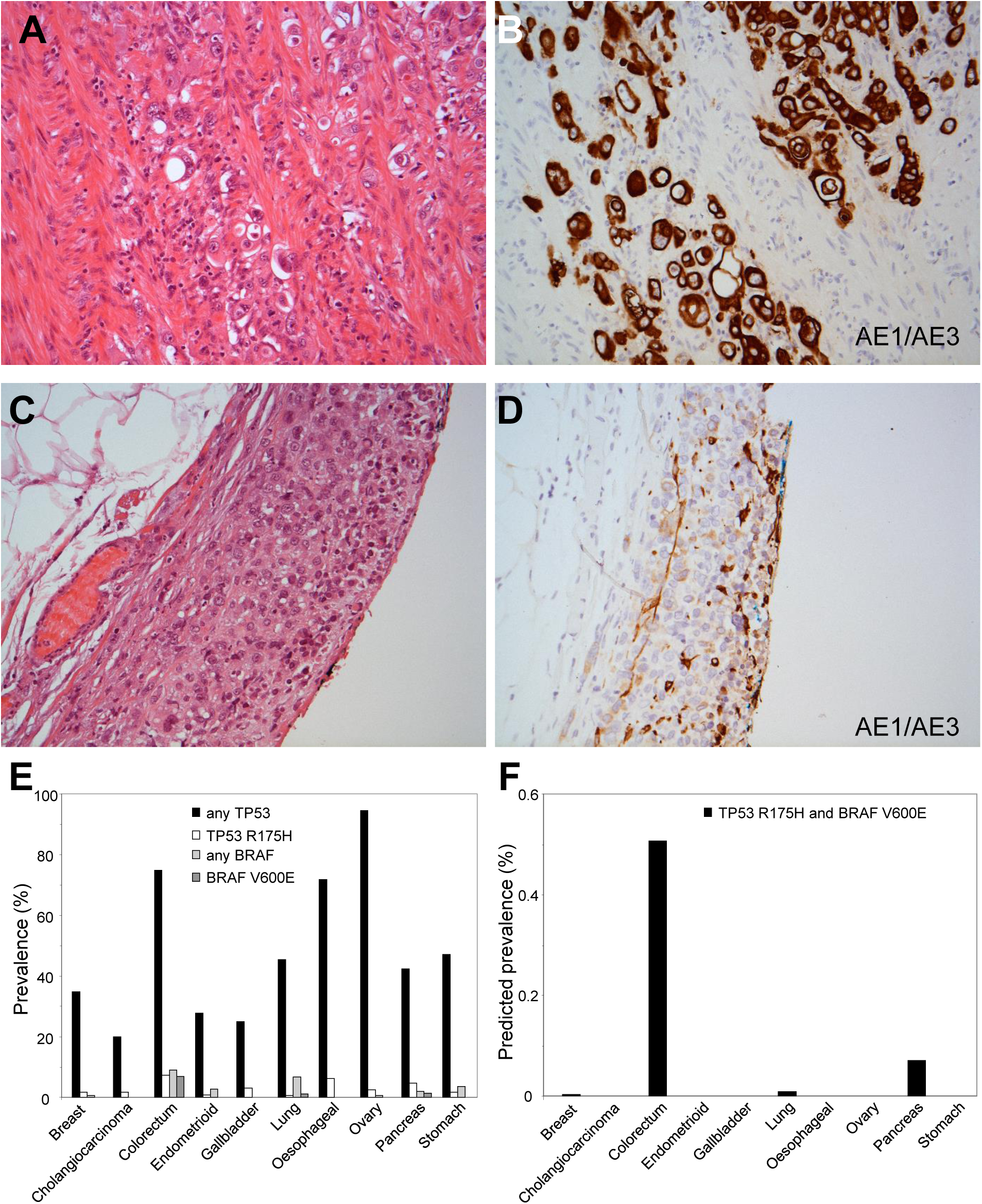
Case study 2. **A, B**. Transverse colon tumor morphology by H&E section (A) and expression of cytokeratin AE1/AE3 by immunohistochemistry (B). **C, D**. Omental tumor morphology by H&E section (C and expression of cytokeratin AE1/AE3 by immunohistochemistry (D). Note expression of AE1/AE3 in benign mesothelial cells but not tumor cells. **E**. Prevalence of all sequence-detected TP53 and BRAF mutations, and specific TP53 (R175H) and BRAF (V600E) mutations in adenocarcinomas from multiple sites. Data from cBioPortal (see text). **F**. Expected prevalence of the co-occurrence of TP53 R175H and BRAF V600E mutations in adenocarcinomas from various sites.

Analysis of cBioPortal data from 3688 adenocarcinomas relevant to females showed that the frequency of TP53 R175H and BRAF V600E were both highest in colorectum, 7.4% and 6.8% respectively (Fig.5E). TP53 R175H and BRAF V600E are predicted to occur in combination in about 0.5% of colorectal carcinomas (if independent, Fig.5F). However, this combination was not found in any of these 3688 adenocarcinomas, including 365 colorectal adenocarcinomas (1.85 tumors were expected in the colorectal set). The low prevalence of TP53 R175H with BRAF V600E combination strongly supported the omental malignancy to be metastatic undifferentiated tumor from the colonic adenocarcinoma.

### Case 3

A male in his 80’s, an ex-smoker, presented with a pleurally-based left lung mass. This was one year after a pT4 N1 trans-glottic poorly differentiated squamous cell carcinoma had been treated by total laryngectomy and radiotherapy. A core biopsy of the lung tumor showed a poorly differentiated non-small cell carcinoma with strong p40 and CK5/6 expression, and weak TTF-1 expression (not shown). Next generation sequencing of TP53, CDKN2A and PIK3CA showed a PIK3CA H1047R mutation (3140A>G) in both the lung and laryngeal tumors. Analysis of mutation data in cBioPortal showed that PIK3CA H1047R mutations were present in only 2.51% SCCs from the head and neck (<1 in 40 tumors) and in 1.12% from lung (about 1 in 89 tumors). PIK3CA H1047R mutations were also uncommon in other p63/p40-positive tumors, present in 1.33% of esophageal SCCs, none of 29 metastatic skin SCCs, and in 0.80% urothelial carcinomas. This data provided strong support that the pleural tumor was a metastasis from the larynx and not a primary lung SCC or a metastasis from an unknown primary.

## DISCUSSION

Metastases are often readily and reliably distinguished from new primary malignancies by a combination of clinical, radiological, morphological and immunohistochemical information. However, sometimes this is not possible, particularly for poorly differentiated tumors. The data presented shows that mutations of specific genes can be helpful to distinguish metastases from a new second malignancy. This appears to be a relatively straightforward analysis, using next generation sequencing methodology and publically available comprehensive tumor mutations data sets. It is possible that this analysis may be improved by using nucleotide rather than amino acid change, and incorporating large scale genomic changes (e.g. amplifications, deletions, translocations, inversions) or DNA modifications such as methylation. Potential limitations could results from poor tumor DNA quality (e.g. sample age, fixation and necrosis), sequencing errors, difficultly classifying variants as pathogenic, mutation-to-wild type reversions and expansion of rare sub-clones that lack truncal mutations.

The case histories illustrate specific situations in which sequence data from a small gene panel clarified conventional pathology information and guided appropriate patient management. In case 1, the solitary lung mass and mediastinal lymphadenopathy suggested primary lung adenocarcinoma. The initial biopsy site was a cytology specimen from one of the lymph nodes, and with current emphasis to limit immunohistochemistry (e.g. TTF-1, p63/p40) in order to preserve material for molecular testing for targeted therapies, this could have resulted in a diagnosis of lung adenocarcinoma. In case 2 the molecular analysis supported a diagnosis of metastatic undifferentiated tumor from the previously identified colonic adenocarcinoma. This was potentially a high stage tumor from an unknown primary, and thus improving the diagnostic certainty potentially limited unnecessary further investigation and therapy. Case 3 illustrates how molecular data can help distinguish metastatic SCC from a primary lung SCC, a common dilemma for pathologists in which morphology and immunohistochemistry are typically unhelpful.

For many tumor types the gene with the highest overall index of diversity was the tumor suppressor TP53 which had both a high prevalence of mutations and a highly diverse array of mutations, as noted previously [45]. Almost all amino acids in TP53 have been found to be mutated in cancers [45]. In contrast, some genes contributed very little to tumor tracking despite being frequently mutated because the mutant subset had a low index of diversity (e.g. BRAF V600E). In this regard KRAS was surprisingly heterogeneous because although many KRAS mutations were codon 12/13 variants, they consisted of multiple amino acid changes occurring at similar frequencies, resulting in a surprisingly high index of diversity. TP53 mutations by themselves were sufficiently diverse that about a third of all tumors had a mutation that would only have 1% likelihood of occurring in a second malignancy. In other words identifying the same TP53 mutation in two different tumors would only occur 1 in 100 times by chance. Therefore, if a germline mutation is excluded, then such data would strongly support the two tumors to be related.

TP53 mutations typically occur early during neoplasia and persist in metastases [46, 47]. Some TP53 mutations may actually promote metastasis [48]. In colorectal adenocarcinoma, mutations in genes other than TP53 are almost always detected in their metastases [11]. This included >99% of the mutations analysed in this study (not shown). Persistence of mutations in metastases is also likely to be generally true for most malignancies as most of these mutations are strong drivers of tumorigenesis.

Diverse mutation profiles were difficult to identify for some tumor types including prostate adenocarcinoma and papillary thyroid carcinoma. However, in clinical practise distinguishing metastatic prostate or thyroid carcinoma from a new primary tumor is usually reliably done with conventional H&E morphology and immunohistochemistry. In contrast distinguishing between metastatic squamous cell carcinoma and a new squamous malignancy is often not possible as the morphology and immunoprofiles of SCCs are often similar. This is not an uncommon clinical dilemma because patients often have risk factors for SCC at several sites (e.g. smoking for oropharyngeal, laryngeal and lung SCC). Compared to adenocarcinomas there are no tissue-of-origin immunostains for SCC, apart from perhaps GATA3 and p16 in specific circumstances. Sequence data can be used to infer aetiology from UV, smoking or HPV, but in contrast to mutation profiling this requires a deeper bioinformatic analysis and does not distinguish between two tumors with the same aetiology.

A potential future approach based upon this study would be similar to that in the case histories, with the modification that the set of genes chosen for sequencing would be those predicted to be the most diverse in the primary malignancy. If the primary malignancy and second tumor have the same mutation profile then the prevalence of the profile in large publically available online data sets such cBioPortal will provide a quantitative estimate of how likely the second tumor is to be a metastasis. Refinement of this method may be to limit the sequencing to the most heterogeneous exons and not whole coding regions, and by avoiding GC-rich sequences. However, the distribution of mutations throughout the TP53 suggests that the entire coding sequence of this gene will remain important to this type of analysis.

## CONCLUSIONS

Most common tumors, including squamous cell carcinomas, have a readily identifiable set of mutations, or mutation profiles that occurs at a sufficiently low prevalence to effectively barcode or fingerprint the tumor. The same mutation profile in the primary tumor and a new lesion provides strong evidence for a metastasis and effectively excludes a new primary malignancy.

## Data Availability

Data is either publicly available in cBioportal.com or where generated on the 3 case studies is available on request as redacted files.

https://www.cbioportal.org/

## DECLARATIONS

## List of abbreviations

FFPE: formalin-fixed paraffin-embedded
FDG-PET: fluorodeoxyglucose-positron emission tomography
CT: computed-tomography
PTC: papillary thyroid carcinoma
SCC: squamous cell carcinoma

## Consent for publication

The data from cases studies were part of standard of care for diagnostic, prognostic and predictive purposes. Research use of this data as shown here was approved by the Peter MacCallum Cancer Centre Ethics committee (project #03/90).

## Availability of data and materials

All data not shown is available on request.

## Authors’ contributions

OWJP conceived the study, analysed and interpreted the data, and wrote the manuscript. Next generation sequencing on the case studies was designed, performed and interpret by RV and AF. PL interpreted data for case study 1 and critically revised the manuscript for intellectual content.

## Acknowledgements

The authors would like to thank the patients, somatic molecular team, Maria Doyle (Research Computing Facility) and the staff of the Department of Anatomical Pathology at the Peter MacCallum Cancer Centre for their contributions to this study. The results shown here are in whole or part based upon data generated by the TCGA Research Network (http://cancergenome.nih.gov/).

## REFERENCES

1. Gao J, Aksoy BA, Dogrusoz U, Dresdner G, Gross B, Sumer SO, Sun Y, Jacobsen A, Sinha R, Larsson E et al: Integrative analysis of complex cancer genomics and clinical profiles using the cBioPortal. Sci Signal 2013, 6(269):pl1.

2. Cerami E, Gao J, Dogrusoz U, Gross BE, Sumer SO, Aksoy BA, Jacobsen A, Byrne CJ, Heuer ML, Larsson E et al: The cBio cancer genomics portal: an open platform for exploring multidimensional cancer genomics data. Cancer Discov 2012, 2(5):401–404.

3. Shah SP, Roth A, Goya R, Oloumi A, Ha G, Zhao Y, Turashvili G, Ding J, Tse K, Haffari G et al: The clonal and mutational evolution spectrum of primary triple-negative breast cancers. Nature 2012, 486(7403):395–399.

4. Banerji S, Cibulskis K, Rangel-Escareno C, Brown KK, Carter SL, Frederick AM, Lawrence MS, Sivachenko AY, Sougnez C, Zou L et al: Sequence analysis of mutations and translocations across breast cancer subtypes. Nature 2012, 486(7403):405–409.

5. Stephens PJ, Tarpey PS, Davies H, Van Loo P, Greenman C, Wedge DC, Nik-Zainal S, Martin S, Varela I, Bignell GR et al: The landscape of cancer genes and mutational processes in breast cancer. Nature 2012, 486(7403):400–404.

6. Ciriello G, Gatza ML, Beck AH, Wilkerson MD, Rhie SK, Pastore A, Zhang H, McLellan M, Yau C, Kandoth C et al: Comprehensive Molecular Portraits of Invasive Lobular Breast Cancer. Cell 2015, 163(2):506–519.

7. Chan-On W, Nairismagi ML, Ong CK, Lim WK, Dima S, Pairojkul C, Lim KH, McPherson JR, Cutcutache I, Heng HL et al: Exome sequencing identifies distinct mutational patterns in liver fluke-related and non-infection-related bile duct cancers. Nature genetics 2013, 45(12):1474–1478.

8. Jiao Y, Pawlik TM, Anders RA, Selaru FM, Streppel MM, Lucas DJ, Niknafs N, Guthrie VB, Maitra A, Argani P et al: Exome sequencing identifies frequent inactivating mutations in BAP1, ARID1A and PBRM1 in intrahepatic cholangiocarcinomas. Nature genetics 2013, 45(12):1470–1473.

9. Seshagiri S, Stawiski EW, Durinck S, Modrusan Z, Storm EE, Conboy CB, Chaudhuri S, Guan Y, Janakiraman V, Jaiswal BS et al: Recurrent R-spondin fusions in colon cancer. Nature 2012, 488(7413):660–664.

10. Comprehensive molecular characterization of human colon and rectal cancer. Nature 2012, 487(7407):330–337.

11. Brannon AR, Vakiani E, Sylvester BE, Scott SN, McDermott G, Shah RH, Kania K, Viale A, Oschwald DM, Vacic V et al: Comparative sequencing analysis reveals high genomic concordance between matched primary and metastatic colorectal cancer lesions. Genome biology 2014, 15(8):454.

12. Kandoth C, Schultz N, Cherniack AD, Akbani R, Liu Y, Shen H, Robertson AG, Pashtan I, Shen R, Benz CC et al: Integrated genomic characterization of endometrial carcinoma. Nature 2013, 497(7447):67–73.

13. Dulak AM, Stojanov P, Peng S, Lawrence MS, Fox C, Stewart C, Bandla S, Imamura Y, Schumacher SE, Shefler E et al: Exome and whole-genome sequencing of esophageal adenocarcinoma identifies recurrent driver events and mutational complexity. Nature genetics 2013, 45(5):478–486.

14. Li M, Zhang Z, Li X, Ye J, Wu X, Tan Z, Liu C, Shen B, Wang XA, Wu W et al: Whole-exome and targeted gene sequencing of gallbladder carcinoma identifies recurrent mutations in the ErbB pathway. Nature genetics 2014, 46(8):872–876.

15. Imielinski M, Berger AH, Hammerman PS, Hernandez B, Pugh TJ, Hodis E, Cho J, Suh J, Capelletti M, Sivachenko A et al: Mapping the hallmarks of lung adenocarcinoma with massively parallel sequencing. Cell 2012, 150(6):1107–1120.

16. Rizvi NA, Hellmann MD, Snyder A, Kvistborg P, Makarov V, Havel JJ, Lee W, Yuan J, Wong P, Ho TS et al: Cancer immunology. Mutational landscape determines sensitivity to PD-1 blockade in non-small cell lung cancer. Science (New York, NY) 2015, 348(6230):124–128.

17. Comprehensive molecular profiling of lung adenocarcinoma. Nature 2014, 511(7511):543–550.

18. Ding L, Getz G, Wheeler DA, Mardis ER, McLellan MD, Cibulskis K, Sougnez C, Greulich H, Muzny DM, Morgan MB et al: Somatic mutations affect key pathways in lung adenocarcinoma. Nature 2008, 455(7216):1069–1075.

19. Integrated genomic analyses of ovarian carcinoma. Nature 2011, 474(7353):609–615.

20. Biankin AV, Waddell N, Kassahn KS, Gingras MC, Muthuswamy LB, Johns AL, Miller DK, Wilson PJ, Patch AM, Wu J et al: Pancreatic cancer genomes reveal aberrations in axon guidance pathway genes. Nature 2012, 491(7424):399–405.

21. Witkiewicz AK, McMillan EA, Balaji U, Baek G, Lin WC, Mansour J, Mollaee M, Wagner KU, Koduru P, Yopp A et al: Whole-exome sequencing of pancreatic cancer defines genetic diversity and therapeutic targets. Nature communications 2015, 6:6744.

22. Barbieri CE, Baca SC, Lawrence MS, Demichelis F, Blattner M, Theurillat JP, White TA, Stojanov P, Van Allen E, Stransky N et al: Exome sequencing identifies recurrent SPOP, FOXA1 and MED12 mutations in prostate cancer. Nature genetics 2012, 44(6):685–689.

23. Taylor BS, Schultz N, Hieronymus H, Gopalan A, Xiao Y, Carver BS, Arora VK, Kaushik P, Cerami E, Reva B et al: Integrative genomic profiling of human prostate cancer. Cancer cell 2010, 18(1):11–22.

24. The Molecular Taxonomy of Primary Prostate Cancer. Cell 2015, 163(4):1011–1025.

25. Grasso CS, Wu YM, Robinson DR, Cao X, Dhanasekaran SM, Khan AP, Quist MJ, Jing X, Lonigro RJ, Brenner JC et al: The mutational landscape of lethal castration-resistant prostate cancer. Nature 2012, 487(7406):239–243.

26. Robinson D, Van Allen EM, Wu YM, Schultz N, Lonigro RJ, Mosquera JM, Montgomery B, Taplin ME, Pritchard CC, Attard G et al: Integrative clinical genomics of advanced prostate cancer. Cell 2015, 161(5):1215–1228.

27. Wang K, Yuen ST, Xu J, Lee SP, Yan HH, Shi ST, Siu HC, Deng S, Chu KM, Law S et al: Whole-genome sequencing and comprehensive molecular profiling identify new driver mutations in gastric cancer. Nature genetics 2014, 46(6):573–582.

28. Comprehensive molecular characterization of gastric adenocarcinoma. Nature 2014, 513(7517):202–209.

29. Kakiuchi M, Nishizawa T, Ueda H, Gotoh K, Tanaka A, Hayashi A, Yamamoto S, Tatsuno K, Katoh H, Watanabe Y et al: Recurrent gain-of-function mutations of RHOA in diffuse-type gastric carcinoma. Nature genetics 2014, 46(6):583–587.

30. Wang K, Kan J, Yuen ST, Shi ST, Chu KM, Law S, Chan TL, Kan Z, Chan AS, Tsui WY et al: Exome sequencing identifies frequent mutation of ARID1A in molecular subtypes of gastric cancer. Nature genetics 2011, 43(12):1219–1223.

31. Stransky N, Egloff AM, Tward AD, Kostic AD, Cibulskis K, Sivachenko A, Kryukov GV, Lawrence MS, Sougnez C, McKenna A et al: The mutational landscape of head and neck squamous cell carcinoma. Science (New York, NY) 2011, 333(6046):1157–1160.

32. Comprehensive genomic characterization of head and neck squamous cell carcinomas. Nature 2015, 517(7536):576–582.

33. Comprehensive genomic characterization of squamous cell lung cancers. Nature 2012, 489(7417):519–525.

34. Song Y, Li L, Ou Y, Gao Z, Li E, Li X, Zhang W, Wang J, Xu L, Zhou Y et al: Identification of genomic alterations in oesophageal squamous cell cancer. Nature 2014, 509(7498):91–95.

35. Lin DC, Hao JJ, Nagata Y, Xu L, Shang L, Meng X, Sato Y, Okuno Y, Varela AM, Ding LW et al: Genomic and molecular characterization of esophageal squamous cell carcinoma. Nature genetics 2014, 46(5):467–473.

36. Li YY, Hanna GJ, Laga AC, Haddad RI, Lorch JH, Hammerman PS: Genomic analysis of metastatic cutaneous squamous cell carcinoma. Clinical cancer research: an official journal of the American Association for Cancer Research 2015, 21(6):1447–1456.

37. Iyer G, Al-Ahmadie H, Schultz N, Hanrahan AJ, Ostrovnaya I, Balar AV, Kim PH, Lin O, Weinhold N, Sander C et al: Prevalence and co-occurrence of actionable genomic alterations in high-grade bladder cancer. Journal of clinical oncology: official journal of the American Society of Clinical Oncology 2013, 31(25):3133–3140.

38. Guo G, Sun X, Chen C, Wu S, Huang P, Li Z, Dean M, Huang Y, Jia W, Zhou Q et al: Whole-genome and whole-exome sequencing of bladder cancer identifies frequent alterations in genes involved in sister chromatid cohesion and segregation. Nature genetics 2013, 45(12):1459–1463.

39. Comprehensive molecular characterization of urothelial bladder carcinoma. Nature 2014, 507(7492):315–322.

40. Van Allen EM, Mouw KW, Kim P, Iyer G, Wagle N, Al-Ahmadie H, Zhu C, Ostrovnaya I, Kryukov GV, O’Connor KW et al: Somatic ERCC2 mutations correlate with cisplatin sensitivity in muscle-invasive urothelial carcinoma. Cancer Discov 2014, 4(10):1140–1153.

41. Hodis E, Watson IR, Kryukov GV, Arold ST, Imielinski M, Theurillat JP, Nickerson E, Auclair D, Li L, Place C et al: A landscape of driver mutations in melanoma. Cell 2012, 150(2):251–263.

42. Krauthammer M, Kong Y, Ha BH, Evans P, Bacchiocchi A, McCusker JP, Cheng E, Davis MJ, Goh G, Choi M et al: Exome sequencing identifies recurrent somatic RAC1 mutations in melanoma. Nature genetics 2012, 44(9):1006–1014.

43. Simpson EH: Measurement of Diversity. Nature 1949(688):688.

44. Hurlbert SH: The Nonconcept of Species Diversity: A Critique and Alternative Parameters. Ecology 1971, 52(4):577–586.

45. Leroy B, Anderson M, Soussi T: TP53 mutations in human cancer: database reassessment and prospects for the next decade. Human mutation 2014, 35(6):672–688.

46. Petitjean A, Mathe E, Kato S, Ishioka C, Tavtigian SV, Hainaut P, Olivier M: Impact of mutant p53 functional properties on TP53 mutation patterns and tumor phenotype: lessons from recent developments in the IARC TP53 database. Human mutation 2007, 28(6):622–629.

47. Chang YL, Wu CT, Shih JY, Lee YC: Comparison of p53 and epidermal growth factor receptor gene status between primary tumors and lymph node metastases in non-small cell lung cancers. Annals of surgical oncology 2011, 18(2):543–550.

48. Yeudall WA: p53 mutation in the genesis of metastasis. Sub-cellular biochemistry 2014, 85:105–117.

